# Effect of Neutrophil Elastase Inhibitor (Sivelestat Sodium) on Oxygenation in Patients with Sepsis Induced Acute Respiratory Distress Syndrome

**DOI:** 10.1101/2024.05.30.24308242

**Authors:** Tiejun Wu, Tao Wang, Jinjiao Jiang, Yue Tang, Lina Zhang, Zhiming Jiang, Fen Liu, Guiqing Kong, Tingfa Zhou, Ruijin Liu, Haipeng Guo, Jie Xiao, Wenqing Sun, Yuye Li, Yingying Zhu, Quan Liu, Weifeng Xie, Yan Qu, Xiaozhi Wang

## Abstract

**Objectives:** Neutrophil elastase (NE) plays an important role in the development of acute respiratory distress syndrome (ARDS). Sivelestat sodium, as a selective NE inhibitor, may improve the outcomes of patients with sepsis-induced ARDS in previous studies, but there is a lack of solid evidence. This trial aimed to evaluate the effect of sivelestat sodium on oxygenation in patients with sepsis-induced ARDS.

**Methods:** We conducted a multicenter, double-blind, randomized, placebo-controlled trial enrolling patients diagnosed with sepsis-induced ARDS admitted within 48 hours of the advent of symptoms. Patients were randomized in a 1:1 fashion to sivelestat or placebo. Trial drugs were administrated as a 24-hour continuous intravenous infusion for a minimum duration of 5 days and a maximum duration of 14 days. The primary outcome was PaO_2_/FiO_2_ ratio improvement on Day5 after randomization, defined by a greater than 50% improvement in PaO_2_/FiO_2_ compared with that on ICU admission or PaO_2_/FiO_2_ reached over 300 mmHg on Day5.

**Results:** The study was stopped midway due to a potential between-group difference in mortality observed during the interim analysis. Overall, a total of 70 patients were randomized, of whom 34 were assigned to receive sivelastat sodium and 36 placebo. On day5, 19/34 (55.9%) patients in the sivelastat group had PaO_2_/FiO_2_ ratio improvement compared with 7/36 (19.4%) patients in the placebo group (risk difference, 0.36; 95% CI, 0.14 to 0.56, *p*<0.001). The Kaplan-Meier curves showed a significantly improved 28-day survival rate in patients receiving sivelestat than those not (hazard ratio, 0.32; 95% CI, 0.11 to 0.95; *p*=0.041).

**Conclusion:** In patients with sepsis-induced ARDS, sivelestat sodium could improve oxygenation within the first five days and may be associated with decreased 28-day mortality.

## Introduction

Sepsis is an aberrant immune response to an infection and a syndrome characterized by organ dysfunction[1]. Lung injury is common in sepsis and acute respiratory distress syndrome (ARDS) is a devastating complication of sepsis[2]. Sepsis is the leading cause of ARDS, accounting for approximately 75% of patients with ARDS[3], and the outcomes of patients with sepsis-induced ARDs is worse than those with ARDS from other causes[4, 5]. However, therapies to prevent or treat sepsis-induced ARDS remain elusive[2].

During the pathogenesis of sepsis-induced ARDS, multiple circulating immune cells are activated and inflammatory mediators are massively released into the circulation, which leads to capillary endothelium injury in the lungs[6, 7]. Following lung injury, immune cells such as neutrophils are recruited to the alveolar space and release large amounts of toxic mediators, including neutrophil elastase(NE)[8, 9]. Previous studies found that systemic inflammatory response syndrome (SIRS) patients with high NE levels were prone to developing ARDS[10] and elevated NE activity was also observed in the bronchoalveolar lavage fluid (BALF) of patients with ARDS[11].

Sivelestat sodium, a small molecule weight, selective and reversible NE inhibitor, was discovered in 1990s[12] and may confer protective effects on pulmonary endothelial injury in sepsis animal models[13–15]. Several clinical studies showed that sivelestat sodium could improve oxygenation, ameliorate lung injury, and reduce the duration of mechanical ventilation in patients with sepsis-induced ARDS[16–18]. However, no causal relationship can be implicated due to the observational nature. Therefore, we conducted a multi-center, randomized controlled study to evaluate the role of sivelestat sodium on oxygenation in patients with sepsis-induced ARDS.

## Materials and methods

### Trial design and oversight

We conducted an investigator-initiated, multi-center, double-blind, randomized, placebo-controlled trial at 12 hospitals across China. The human research ethics committee at each hospital approved the protocol. Patients or their surrogates provided written informed consent before enrollment. This trial was designed by the authors, who collected and analyzed the data, vouched for the accuracy and completeness of the data and the adherence of the trial to the protocol, wrote and agreed on the submission of the manuscript. Shanghai Huilun (Jiangsu) Pharmaceutical Co., Ltd. supplied the trial drugs but had no role in designing or conducting the trial, or analysing the data and did not have access to the data before publication. The trial was registered on the Chinese Clinical Trial Registry (ChiCTR2200056892) before enrollment began.

### Study population

Patients diagnosed with sepsis aged between 18 to 75 years old and with ARDS admitted to any of the participating sites within 48 hours of ARDS onset were eligible for inclusion. The diagnosis of sepsis was according to sepsis 3.0 criteria[19] and diagnosis of ARDS was based on Berlin criteria[20]. Patients were excluded if they were pregnant or lactating women, diagnosed with neutropenia, received chemotherapeutic agents or immunomodulatory drugs or high-dose corticosteroid therapy for more than 5 days, with a known history of severe cardiovascular, respiratory, renal, or hepatic diseases, or had disseminated intravascular coagulation, end-stage malignancy, mental illness, etc. Detailed inclusion and exclusion criteria were provided in the Supplementary Protocol.

### Randomization, blinding and interventions

Each eligible participant was assigned randomly from a computer-generated sequence to either the sivelestat sodium or placebo group in a 1:1 ratio, using a block size of 4 stratified by site. The random allocation sequence was generated by a third party independent of the study. Allocation concealment was achieved using blinded medication packs. Participants, data collectors, and investigators assessing outcome data will be blinded to the treatment allocation.

Patients were assigned to receive a 24-hour continuous intravenous infusion of sivelastat sodium at a rate of 0.2 mg/kg/h, for a minimum duration of 5 days and a maximum duration of 14 days or a placebo during the same study period. All other treatments were administered at the discretion of the treating clinicians.

### Trial outcomes

The primary outcome was PaO_2_/FiO_2_ ratio improvement on Day5 after randomization. PaO_2_/FiO_2_ ratio improvement on Day5 was defined as a greater than 50% improvement in PaO_2_/FiO_2_ compared with that on ICU admission or PaO_2_/FiO_2_ reached over 300 mmHg on Day5. Secondary outcomes included PaO_2_/FiO_2_ ratio on Day3, 5, 7 and 28-day mortality, ventilator free days (28-VFDs) with 28 days, ICU and hospital free days within 28 days.

### Sample size estimation

Based on previous studies[16, 17], it is estimated that 35% of the study patients in the control group would reach the primary endpoint (oxygenation index improvement on Day5). We estimated that a sample size of 122 participants could provide 80% power at a two-sided alpha level of 0.05 to detect an absolute 25% elevation in the primary endpoint from the sivelestat sodium intervention. The calculation was implemented using the PASS 11.0 software (PASS, NCSS software, Kaysville, USA). In this trial, we plan to randomize 142 patients in total (71 per group) after allowing for a less than 15% withdrawal. This study employed one planned interim analysis that was conducted by an independent Data and Safety Monitoring Board (DSMB) after the first 70 participants enrolled. This analysis would have led to a recommendation to stop the trial if concerns about participant safety had been raised.

### Statistical analysis

Continuous data were reported as means and standard deviations (SD) when normally distributed or as medians and interquartile ranges (IQR) when not normally distributed. The normality of continuous variables will be examined using the Shapiro–Wilk test. Categorical data will be expressed as numbers and percentages.

The primary analysis was based on the intention-to-treat (ITT) population, defined as all enrolled patients from the participating sites. We used generalized linear model (GLM) to compare the difference in the primary outcome (PaO_2_/FiO_2_ ratio improvement on Day5) between groups. We analyzed secondary outcomes also using GLM. Risk differences and its 95% confidence interval (CI) were calculated for categorical variables, and mean differences (95% CI) for continuous variables. Kaplan–Meier curves were used to compare the 28-day survival curves after randomization. The difference between two-groups was calculated by tested by log-rank test and its hazard ratio (HR) and 95%CIs was calculated by Mantel-Cox regression model.

Four pre-defined subgroup analyses were conducted for the primary endpoint according to (1) age (dichotomized at 50 years old), (2) APACHE II score at enrollment (dichotomized at 15), (3) septic shock at enrollment, and (4) PaO_2_/FiO_2_ ratio at enrollment (dichotomized at 200). Adverse event analyses were reported for all the participants who received the study treatment. Analyses were conducted using R 4.2.3 software. Statistical tests were two-sided, and p values < 0.05 were considered statistically significant.

## Results

### Recruitment and baseline characteristics

During the study period, 282 patients with sepsis combined with ARDS were assessed for eligibility, of whom 70 were enrolled in the trial from nine hospitals across China. The trial recruitment was halted by the DSMB after the interim analysis owing to observed between-group difference in mortality, and the DSMB requested to unblind the data. After reviewing the unblinded data, the DSMB concluded that the trial should be stopped midway due to potential mortality benefits from the trial intervention, and the trial was then formally stopped. The numbers of cases from each site were shown in online Supplemental table 1. Among those 70 randomized patients, 34 were assigned to receive sivelastat sodium and 36 placebo. All randomized participants completed follow-up and were included in the primary analysis (**Figure 1**).

**Figure 1.**
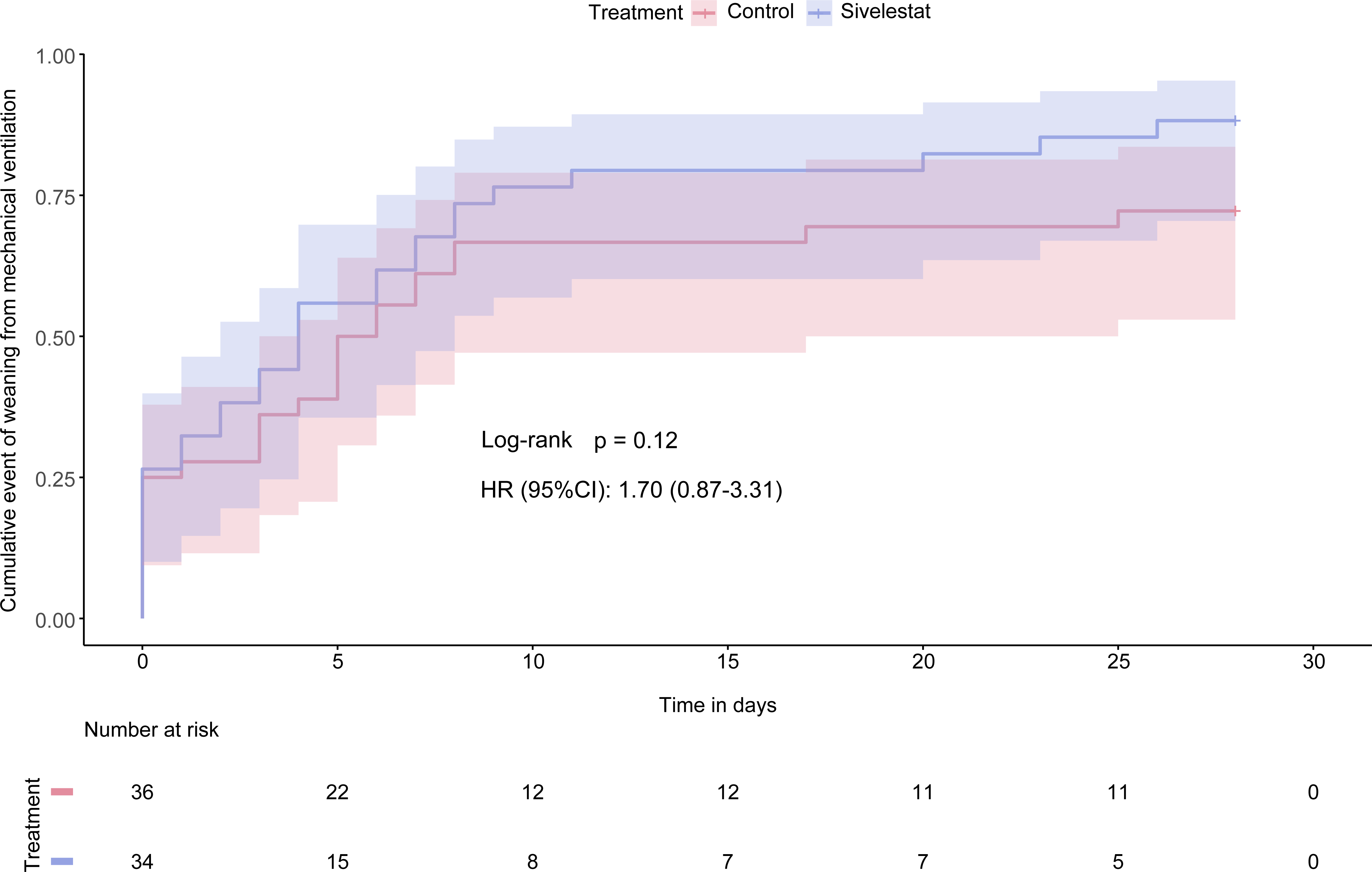
Study flowchart.

The characteristics of the participants at baseline were evenly distributed between the two trial groups (**Table 1**). The majority of the trial participants required mechanical ventilation (52/70, 74.3%) at admission. The median (IQR) PaO_2_/FiO_2_ ratio was 136.0 (104.2-163.0) mmHg in the sivelastat group and 161.0 (144.0-195.0) mmHg in the placebo group.

**Table 1.** Baseline characteristics of the study subjects.

| Characteristics | Sivelestat group<br>(N=34) | Placebo group<br>(N=36) |
| --- | --- | --- |
| Mean Age±SD, yr | 61.2±11.4 | 56.5±17.6 |
| Gender, n (%) |  |  |
| Women | 12 (35.3) | 13 (36.1) |
| Men | 22 (64.7) | 23 (63.9) |
| Mean BMI±SD, kg/m<sup>2</sup> | 25.0±4.4 | 25.5±3.6 |
| Median Charlson score (IQR) | 4.0 (2.5-5.0) | 5.0 (2.5-5.5) |
| Comorbidities, no. (%) |  |  |
| Hypertension | 15 (44.1) | 11 (30.6) |
| Diabetes mellitus | 11 (32.4) | 4 (11.1) |
| Coronary heart disease | 6 (17.6) | 4 (11.1) |
| History of covid-19 infection, n (%) | 4 (11.8) | 8 (22.2) |
| Use of mechanical ventilation, n (%) | 25 (73.5) | 27 (75.0) |
| Ventilator related parameters, median (IQR) |  |  |
| Lung compliance, ml/cmH<sub>2</sub>O | 51.0 (30.8-58.5) | 39.3 (33.2-48.4) |
| Positive end-expiratory pressure at randomization, cmH<sub>2</sub>O | 5 (5-8) | 5 (5-8) |
| Clinical parameters |  |  |
| Presence of sepsis, no. (%) | 34 (100.0) | 35 (97.2) |
| Presence of septic shock, no. (%) | 16 (47.1) | 15 (41.7) |
| Median APACHE II score (IQR) | 20.5 (12.0-25.0) | 17.5 (11.3-24.0) |
| Median SOFA score (IQR) | 7.0 (4.8-10.0) | 6.0 (4.3-9.8) |
| Median lac (IQR), mmol/L | 1.2 (1.0-2.8) | 1.4 (1.0-1.8) |
| Median PaO<sub>2</sub>/FiO<sub>2</sub> ratio (IQR) | 136.0 (104.2-163.0) | 161.0 (144.0-195.0) |
| Laboratory parameters, median (IQR) |  |  |
| Serum NE concentration, pg/ml, $\times 10^4$ | 31.0 (7.8-35.5) | 32.7 (15.6-37.7) |

### Primary and secondary outcomes

On day5 after randomization, 19/34 (55.9%) patients in the sivelastat group had PaO_2_/FiO_2_ ratio improvement compared with 7/36 (19.4%) patients in the placebo group (risk difference, 0.36; 95% CI, 0.14 to 0.56, *p*<0.001). In addition, the PaO_2_/FiO_2_ ratio constantly differ between groups on day3, day5 and day7 (**Table 2**).

**Table 2.**
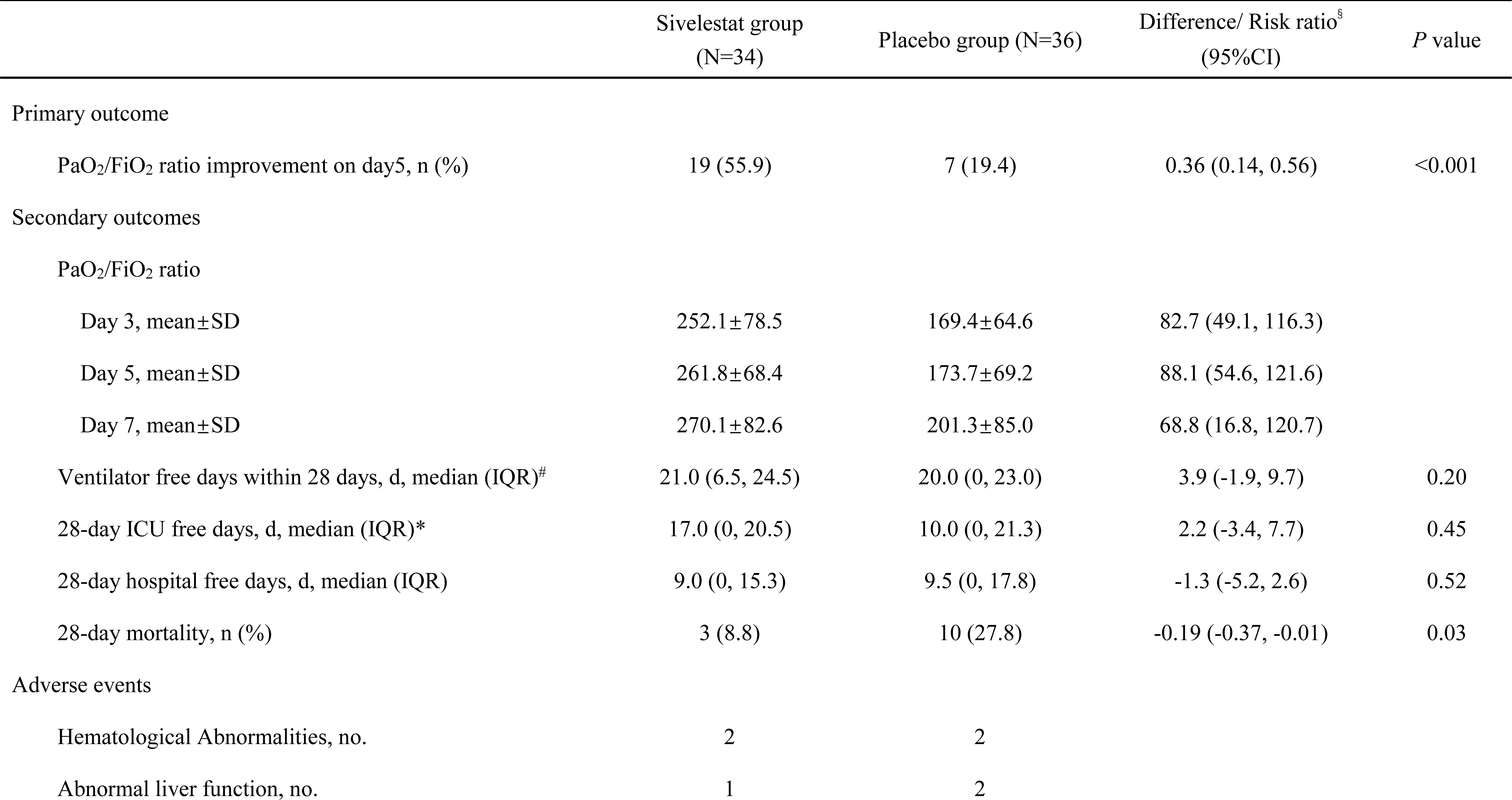
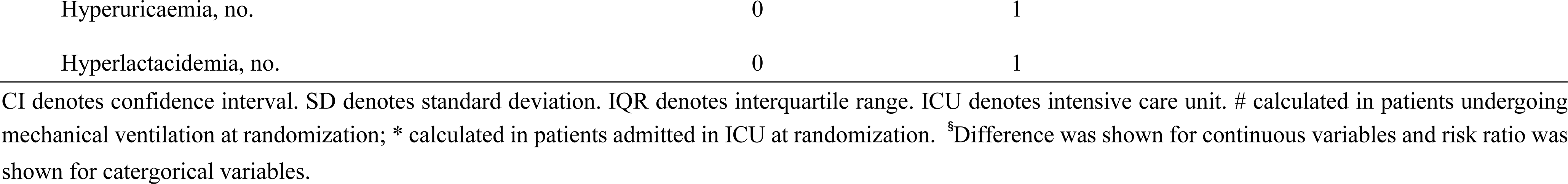
Trial outcomes. CI denotes confidence interval. SD denotes standard deviation. IQR denotes interquartile range. ICU denotes intensive care unit. # calculated in patients undergoing mechanical ventilation at randomization; *calculated in patients admitted in ICU at randomization.

|  | Sivelestat group<br>(N=34) | Placebo group (N=36) | Difference/ Risk ratio<sup>§</sup><br>(95% CI) | *P* value |
| --- | --- | --- | --- | --- |
| Primary outcome |  |  |  |  |
| PaO<sub>2</sub>/FiO<sub>2</sub> ratio improvement on day5, n (%) | 19 (55.9) | 7 (19.4) | 0.36 (0.14, 0.56) | <0.001 |
| Secondary outcomes |  |  |  |  |
| PaO<sub>2</sub>/FiO<sub>2</sub> ratio |  |  |  |  |
| Day 3, mean±SD | 252.1±78.5 | 169.4±64.6 | 82.7 (49.1, 116.3) |  |
| Day 5, mean±SD | 261.8±68.4 | 173.7±69.2 | 88.1 (54.6, 121.6) |  |
| Day 7, mean±SD | 270.1±82.6 | 201.3±85.0 | 68.8 (16.8, 120.7) |  |
| Ventilator free days within 28 days, d, median (IQR)<sup>#</sup> | 21.0 (6.5, 24.5) | 20.0 (0, 23.0) | 3.9 (-1.9, 9.7) | 0.20 |
| 28-day ICU free days, d, median (IQR)* | 17.0 (0, 20.5) | 10.0 (0, 21.3) | 2.2 (-3.4, 7.7) | 0.45 |
| 28-day hospital free days, d, median (IQR) | 9.0 (0, 15.3) | 9.5 (0, 17.8) | -1.3 (-5.2, 2.6) | 0.52 |
| 28-day mortality, n (%) | 3 (8.8) | 10 (27.8) | -0.19 (-0.37, -0.01) | 0.03 |
| Adverse events |  |  |  |  |
| Hematological Abnormalities, no. | 2 | 2 |  |  |
| Abnormal liver function, no. | 1 | 2 |  |  |

|  |  |  |
| --- | --- | --- |
| Hyperuricaemia, no. | 0 | 1 |
| Hyperlactacidemia, no. | 0 | 1 |

CI denotes confidence interval. SD denotes standard deviation. IQR denotes interquartile range. ICU denotes intensive care unit. # calculated in patients undergoing mechanical ventilation at randomization; \* calculated in patients admitted in ICU at randomization. <sup>§</sup>Difference was shown for continuous variables and risk ratio was shown for categorical variables.

Patients in the sivelastat group had a median of 21.0 VFDs (IQR, 6.5-24.5) within the first 28 days compared with 20.0 VFDs (IQR, 0-23.0) for those receiving placebo. The mean difference in VFDs between groups was 3.9 days (95% CI, −1.9 to 9.7, *p*=0.20). No significant difference in cumulative event of weaning from mechanical ventilation between treatment groups was observed (hazard ratio, 1.70; 95% CI, 0.87 to 3.31, p=0.12, supplementary figure 1). The 28-day mortality was 3/34 (8.8%) in patients receiving sivelastat and 10/36 (27.8%) in those receiving placebo (risk difference, −0.19; 95% CI, −0.37 to −0.01, *p*=0.03). The Kaplan-Meier curves showed a significantly improved survival rate in patients receiving sivelestat than those not (HR, 0.32; 95% CI, 0.11 to 0.95; log-rank *p*=0.041) (**Figure 2**). The ICU and hospital free days within 28 days were both comparable between two groups (**Table 2**).

**Figure 2.**
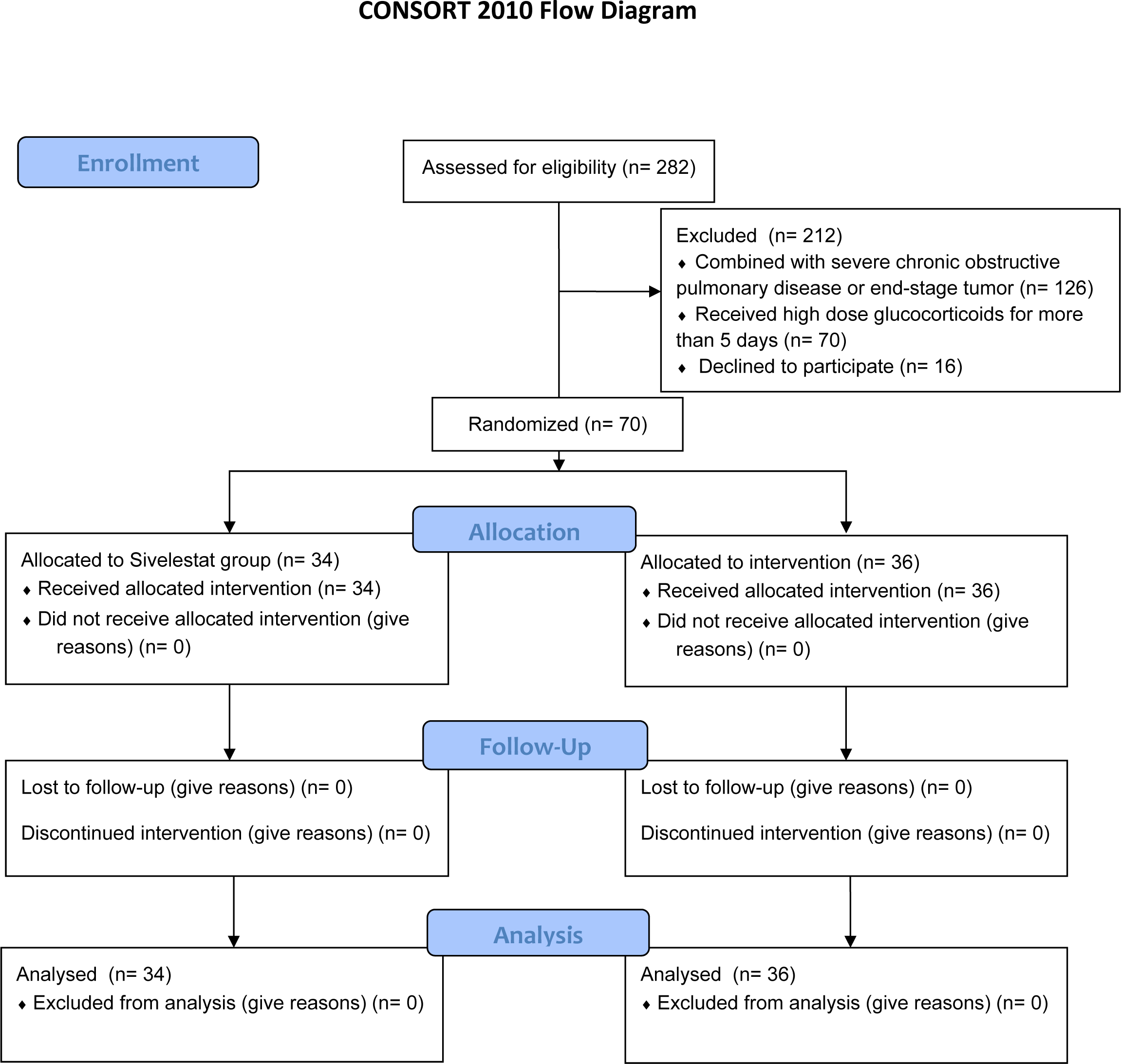
Survival curve. HR denotes hazard ratio. CI denotes confidence interval.

### Subgroup analysis

Subgroup analysis suggested the treatment effect of sivelestat on the primary outcome had trends toward more significant in patients with age≥50, with APACHE II score<15, without septic shock and with PaO_2_/FiO_2_ ratio≥200 at enrollment (**Figure 3**).

**Figure 3.**
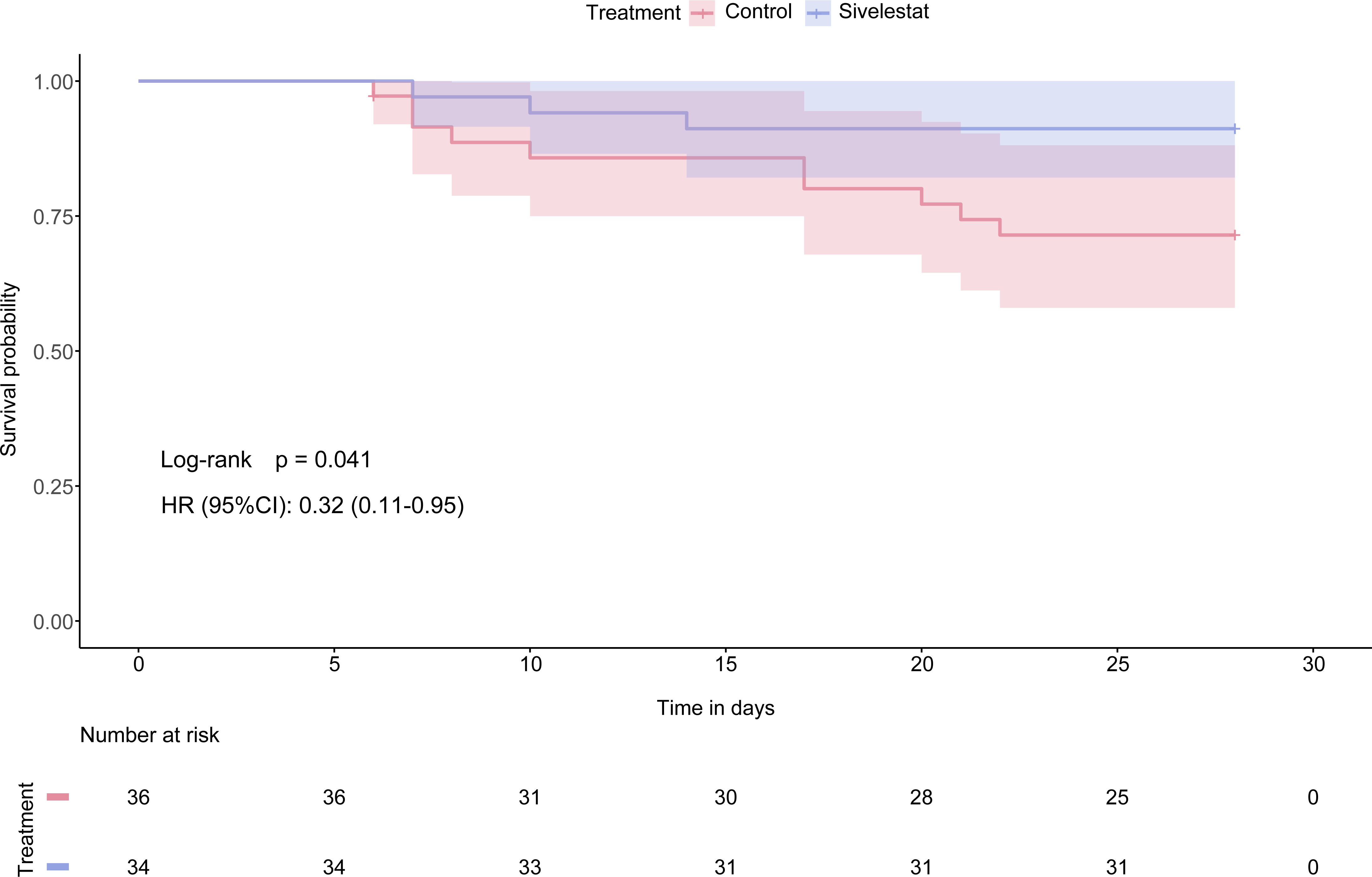
Subgroup analysis for the primary outcome. RD denotes risk difference. CI denotes confidence interval. APACHE II denotes AcutePhysiology and Chronic Health Evaluation II.

### Adverse events

The number of adverse events did not differ meaningfully between the trial groups. Details regarding adverse events are provided in **Table 2**.

## Disscussion

In this multicenter, double-blind, randomized, placebo-controlled trial, the use of sivelestat sodium could improve oxygenation within the first week in patients with sepsis-induced ARDS. Moreover, it was associated with decreased 28-day mortality, though there is no difference in ventilator-free days or other outcomes. Subgroup analysis showed that age, the baseline respiratory function, disease severity and septic status may affect the efficacy of sivelestat sodium, favouring sivelestat use in patients with age≥50, with APACHE II score<15, with PaO_2_/FiO_2_ ratio≥200, and without septic shock.

Two large clinical trials demonstrated discordant effects of sivelestat sodium in patients with acute lung injury (ALI)[21, 22]. The phase III Japanese study by Tamakuma et al. included 230 ALI/ARDS patients combined with SIRS, and sivelestat was shown to increase pulmonary function, reduce duration of mechanical ventilation, and shorten ICU stay[21]. An international multicenter double-blind, placebo-controlled Phase II study (STRIVE study) randomized 492 mechanically ventilated patients with ALI/ARDS[22], and the results showed that sivelestat did not change 28-day mortality or VFDs. Furthermore, a negative trend in long-term 180-day mortality rate was noted, and the trial was then stopped midway per the recommendation from the DSMB.

The discrepancy between the two studies may be due to differences in the characteristics of study patients, such as age, baseline respiratory function, disease severity and septic status. The patients enrolled in the phase III Japanese study had a narrower age distribution and better respiratory function than those in the STRIVE study. In addition, clinical studies reporting positive results with sivelestat therapy had mainly enrolled ARDS patients with a Lung Injury Score <2.5, whereas the majority of the patients in the STRIVE study had a Lung Injury Score >2.5[23, 24]. A post-hoc analysis of the STRIVE patients involving those who had a mean Lung Injury Score≤2.5, revealed favourable trends in mortality and VFDs in patients receiving sivelestat[22]. On the other hand, it is suggested that the different proportions of septic patients may have contributed to the discordant results among these studies (58% vs. 69%). Our results were consistent with the above findings, showing that sivelestat may confer larger treatment effects in patients with sepsis-induced ARDS, especially in patients with less severe disease, including those with APACHE II score<15 or with PaO_2_/FiO_2_ ratio≥200 or without septic shock. Taken together, our study suggests that sivelestat could be effective in patients with mild sepsis-induced ARDS, and may be associated with survival benefits in this popualtion.

The above results can also be explained from a pathophysiological point of view. Neutrophil activation and NE release are very early biological events in the pathogenesis of ARDS[25]. Previous research showed a significant increase in blood NE in patients with sepsis-induced ARDS[26] and a decrease in blood NE after sivelestat administration[27, 28], suggesting that the therapeutic effect of sivelestat is related to the inhibition of NE. Recent studies have shown that damage to the endothelial glycocalyx is a critical factor in the development and progression of ARDS[29, 30]. In addition, our preclinical research has shown that sivelestat can reduce endothelial glycocalyx damage by inhibiting the production of NETs, improve endothelial cell permeability, attenuate lung histopathological injury and ultimately improve survival in sepsis-induced ALI model mice. Further molecular docking and visualisation analysis showed that sivelestat could bind with high affinity to the key ferroptosis protein glutathione peroxidase (GPX4), increase the expression of GPX4 and thus interfere with the process of ferroptosis[26]. Therefore, sivelestat may have pleiotropic effects on ARDS and may not be limited to interfering with NE.

The trial had several limitations. First, the current sample size was not powered to detect mortality difference, and stopping the trial midway further weaken the robustness of the results. Thus, the results of this trial should be interpreted cautiously. Second, subjective factors contribute to the decision to wean patients from mechanical ventilation, which may bias the VFDs. Third, the results could be strengthened had more inflammatory mediators and other surrogate measurements, including IL-6, IL-8, IL-10, TNF-IZI been measured. Last, we did not observe the effects of sivelestat on long-term outcomes, like pulmonary function after hospital discharge.

## Conclusion

In patients with sepsis-induced ARDS, sivelestat sodium could improve oxygenation within the first week, and was associated with decreased 28-day mortality, particualrly in patients with less severe disease, including those with APACHE II score<15 or with PaO_2_/FiO_2_ ratio≥200 or without septic shock. Further large-scale RCTs are needed to confirm the effects of sivelestat on mortality in this population.

## Data Availability

All data produced in the present work are contained in the manuscript.

## Ethics and approval statement

The protocol and the informed consent document have been reviewed and approved by the Institutional Ethics Committee of all participating centers. Study investigators will provide potential participants with verbal and written information prior to inclusion in the study. Informed consent will be provided from participants or their authorized representatives. The study was registered in the Clinical Trials Register (ChiCTR2200056892).

## Funding

This study is supported by Shanghai Huilun (Jiangsu) Pharmaceutical Co., Ltd., which is also responsible for the supply of the study drug andplacebo as well as distribution to the participating centers. Funding agencyhad no input into the design, conduct, data collection, statistical analysis, orwriting of the manuscript.

## Authorship contribution statement

Tiejun Wu, Tao Wang and Xiaozhi Wang: drafting of the manuscript; material or technique support; critical revision of the manuscript. Jinjiao Jiang, Yue Tang, Lina Zhang, Zhiming Jiang, Fen Liu, Guiqing Kong, Tingfa Zhou, Ruijin Liu, Haipeng Guo, Jie Xiao, Wenqing Sun, Yuye Li, Yingying Zhu, Quan Liu, Weifeng Xie, Yan Qu: Patient screening and data collection. Xiaozhi Wang: analysis and interpretation of data. Tiejun Wu and Tao Wang: conceived, designed and supervised the study; analysis and interpretation of data; critical revision of the manuscript; obtained funding. All authors have read the manuscript and approved its submission.

## Declaration of competing interest

The authors have declared that no competing interest exists.

---

**Table S1.**
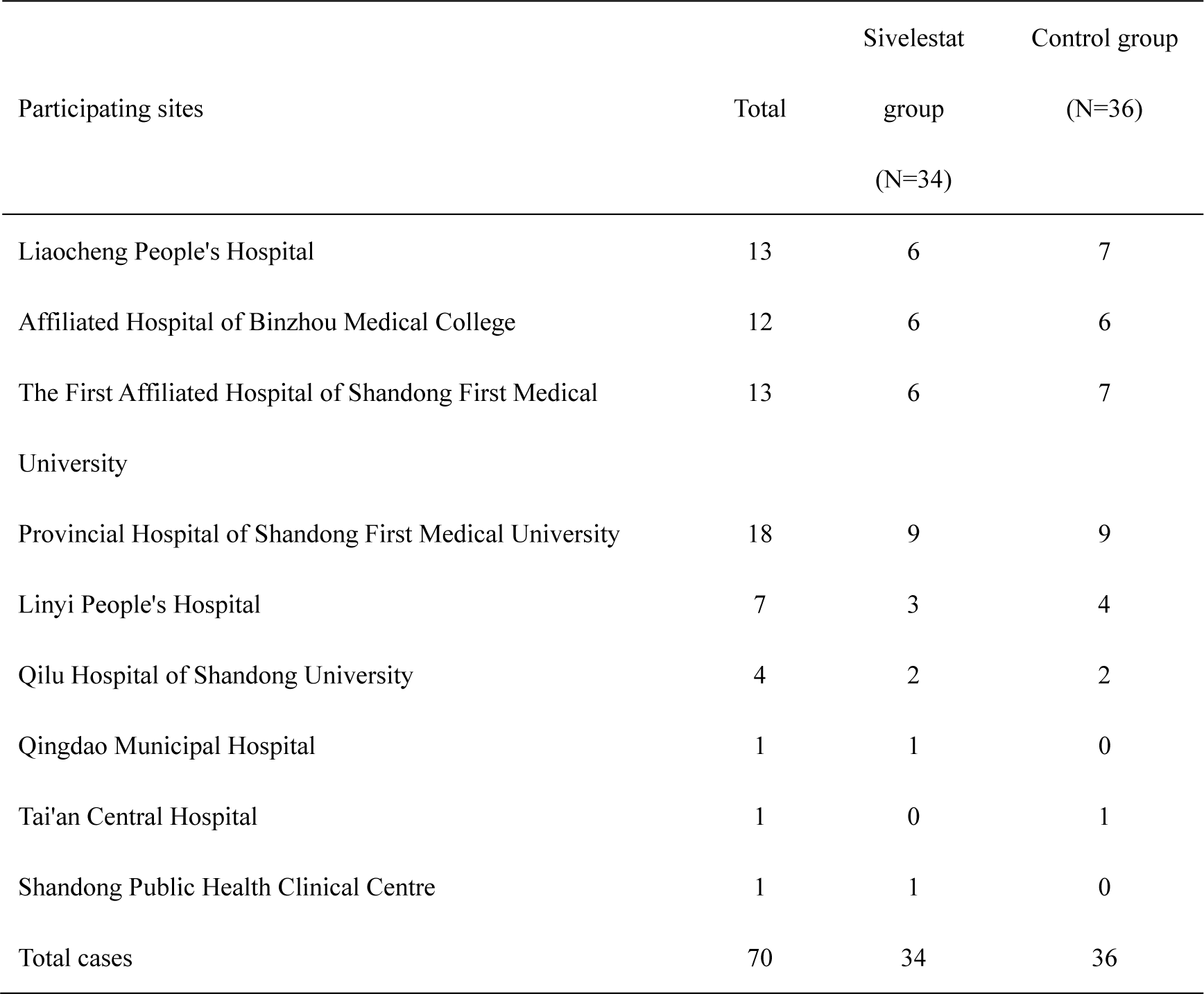
Numbers of cases from each site.

**Figure S1.**
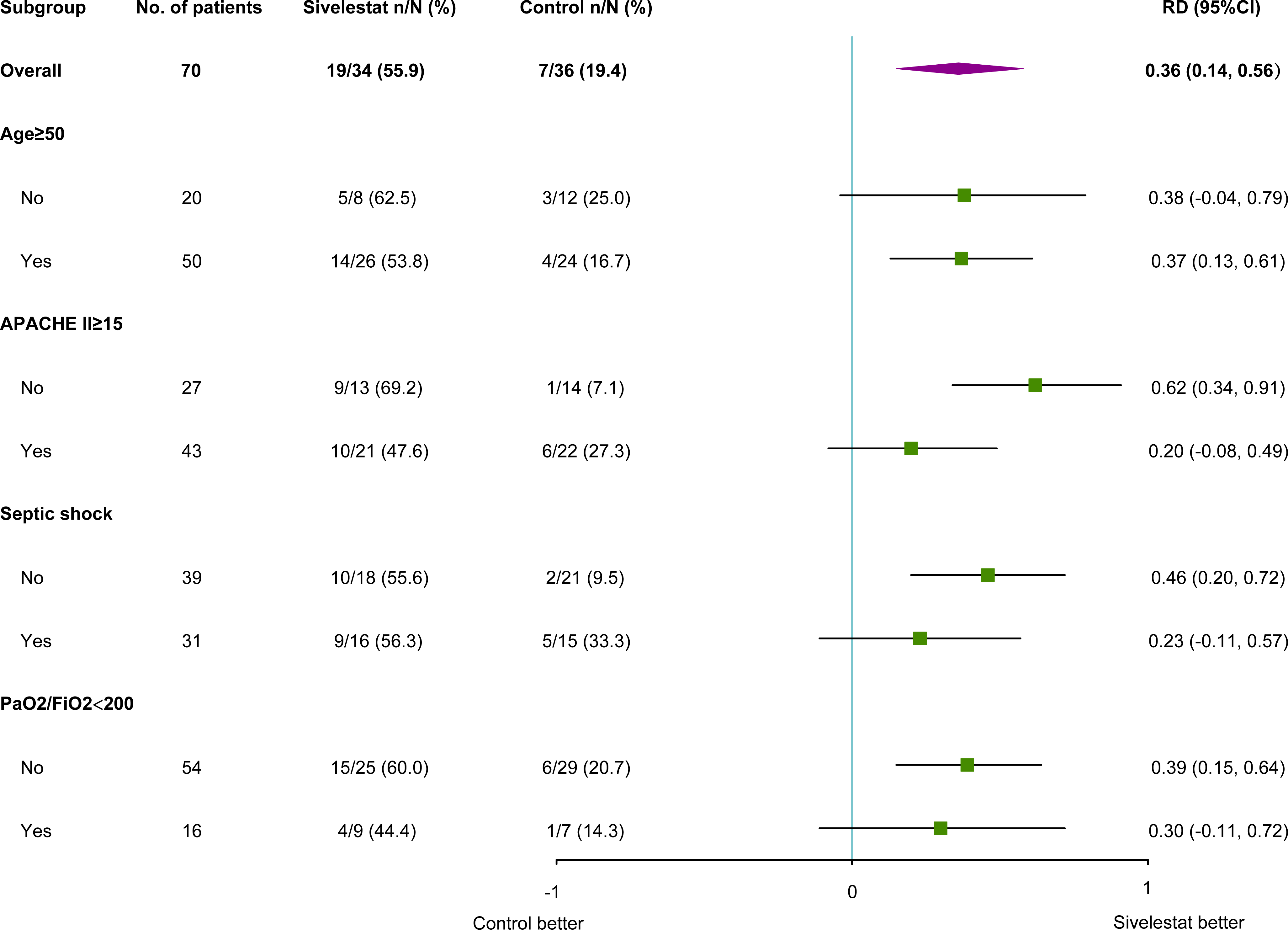
Cumulative event of weaning from mechanical ventilation. HR denotes hazard ratio. CI denotes confidence interval.

Ethical apprval.

**Ethical apprval**
| Organization | Ethical<br>state | Number (no.) | Date |
| --- | --- | --- | --- |
| Liaocheng People's Hospital | Approve | 2022021 | 2022-02-15 |
| Affiliated Hospital of Binzhou Medical College | Approve | 2022-015-1 | 2022-03-10 |
| Provincial Hospital of Shandong First Medical University | Approve | 2022-144 | 2022-04-13 |
| The First Affiliated Hospital of Shandong First Medical University | Approve | YXLL-KY-2022(046) | 2022-06-01 |
| Linyi People's Hospital | Approve | YX200375 | 2022-04-29 |
| Qilu Hospital of Shandong University | Approve | KYLL-2022B-014 | 2022-04-24 |
| Shandong Public Health Clinical Centre | Approve | GWLCZXEC<br>2022-84 | 2022-09-13 |
| Tai'an Central Hospital | Approve | 2022-06-22 | 2022-06-22 |
| Qingdao Municipal Hospital | Approve | 2022-06-30 | 2022-06-30 |

## Notes

### Competing Interest Statement

The authors have declared no competing interest.

### Clinical Trial

ChiCTR2200056892

### Author Declarations

(1)Ethics committee/IRB of Liaocheng People's Hospital gave ethical approval for this work. (2)Ethics committee/IRB of Affiliated Hospital of Binzhou Medical College gave ethical approval for this work. (3)Ethics committee/IRB of Provincial Hospital of Shandong First Medical University gave ethical approval for this work. (4)Ethics committee/IRB of The First Affiliated Hospital of Shandong First Medical University gave ethical approval for this work. (5)Ethics committee/IRB of Linyi People's Hospital gave ethical approval for this work. (6)Ethics committee/IRB of Qilu Hospital of Shandong University gave ethical approval for this work. (7)Ethics committee/IRB of Shandong Public Health Clinical Centre gave ethical approval for this work. (8)Ethics committee/IRB of Tai'an Central Hospital gave ethical approval for this work. (9)Ethics committee/IRB of Qingdao Municipal Hospital gave ethical approval for this work.

## Reference

1. Singer M, Deutschman CS, Seymour CW, Shankar-Hari M, Annane D, Bauer M, Bellomo R, Bernard GR, Chiche JD, Coopersmith CM et al: The Third International Consensus Definitions for Sepsis and Septic Shock (Sepsis-3). Jama 2016, 315(8):801–810.

2. Beitler JR, Thompson BT, Baron RM, Bastarache JA, Denlinger LC, Esserman L, Gong MN, LaVange LM, Lewis RJ, Marshall JC et al: Advancing precision medicine for acute respiratory distress syndrome. Lancet Respir Med 2022, 10(1):107–120.

3. Bellani G, Laffey JG, Pham T, Fan E, Brochard L, Esteban A, Gattinoni L, van Haren F, Larsson A, McAuley DF et al: Epidemiology, Patterns of Care, and Mortality for Patients With Acute Respiratory Distress Syndrome in Intensive Care Units in 50 Countries. Jama 2016, 315(8):788–800.

4. Stapleton RD, Wang BM, Hudson LD, Rubenfeld GD, Caldwell ES, Steinberg KP: Causes and timing of death in patients with ARDS. Chest 2005, 128(2):525–532.

5. Sheu CC, Gong MN, Zhai R, Chen F, Bajwa EK, Clardy PF, Gallagher DC, Thompson BT, Christiani DC: Clinical characteristics and outcomes of sepsis-related vs non-sepsis-related ARDS. Chest 2010, 138(3):559–567.

6. Englert JA, Bobba C, Baron RM: Integrating molecular pathogenesis and clinical translation in sepsis-induced acute respiratory distress syndrome. JCI Insight 2019, 4(2).

7. Xu H, Sheng S, Luo W, Xu X, Zhang Z: Acute respiratory distress syndrome heterogeneity and the septic ARDS subgroup. Front Immunol 2023, 14:1277161.

8. Zeiher BG, Matsuoka S, Kawabata K, Repine JE: Neutrophil elastase and acute lung injury: prospects for sivelestat and other neutrophil elastase inhibitors as therapeutics. Crit Care Med 2002, 30(5 Suppl):S281–287.

9. Sahebnasagh A, Saghafi F, Safdari M, Khataminia M, Sadremomtaz A, Talaei Z, Rezai Ghaleno H, Bagheri M, Habtemariam S, Avan R: Neutrophil elastase inhibitor (sivelestat) may be a promising therapeutic option for management of acute lung injury/acute respiratory distress syndrome or disseminated intravascular coagulation in COVID-19. J Clin Pharm Ther 2020, 45(6):1515–1519.

10. Kodama T, Yukioka H, Kato T, Kato N, Hato F, Kitagawa S: Neutrophil elastase as a predicting factor for development of acute lung injury. Intern Med 2007, 46(11):699–704.

11. McGuire WW, Spragg RG, Cohen AB, Cochrane CG: Studies on the pathogenesis of the adult respiratory distress syndrome. J Clin Invest 1982, 69(3):543–553.

12. Kawabata K, Suzuki M, Sugitani M, Imaki K, Toda M, Miyamoto T: ONO-5046, a novel inhibitor of human neutrophil elastase. Biochem Biophys Res Commun 1991, 177(2):814–820.

13. Suzuki K, Okada H, Takemura G, Takada C, Kuroda A, Yano H, Zaikokuji R, Morishita K, Tomita H, Oda K et al: Neutrophil Elastase Damages the Pulmonary Endothelial Glycocalyx in Lipopolysaccharide-Induced Experimental Endotoxemia. Am J Pathol 2019, 189(8):1526–1535.

14. Suda K, Takeuchi H, Hagiwara T, Miyasho T, Okamoto M, Kawasako K, Yamada S, Suganuma K, Wada N, Saikawa Y et al: Neutrophil elastase inhibitor improves survival of rats with clinically relevant sepsis. Shock 2010, 33(5):526–531.

15. Hagiwara S, Iwasaka H, Togo K, Noguchi T: A neutrophil elastase inhibitor, sivelestat, reduces lung injury following endotoxin-induced shock in rats by inhibiting HMGB1. Inflammation 2008, 31(4):227–234.

16. Gao X, Zhang R, Lei Z, Guo X, Yang Y, Tian J, Huang L: Efficacy, safety, and pharmacoeconomics of sivelestat sodium in the treatment of septic acute respiratory distress syndrome: a retrospective cohort study. Ann Palliat Med 2021, 10(11):11910–11917.

17. Hayakawa M, Katabami K, Wada T, Sugano M, Hoshino H, Sawamura A, Gando S: Sivelestat (selective neutrophil elastase inhibitor) improves the mortality rate of sepsis associated with both acute respiratory distress syndrome and disseminated intravascular coagulation patients. Shock 2010, 33(1):14–18.

18. Miyoshi S, Hamada H, Ito R, Katayama H, Irifune K, Suwaki T, Nakanishi N, Kanematsu T, Dote K, Aibiki M et al: Usefulness of a selective neutrophil elastase inhibitor, sivelestat, in acute lung injury patients with sepsis. Drug Des Devel Ther 2013, 7:305–316.

19. Shankar-Hari M, Phillips GS, Levy ML, Seymour CW, Liu VX, Deutschman CS, Angus DC, Rubenfeld GD, Singer M: Developing a New Definition and Assessing New Clinical Criteria for Septic Shock: For the Third International Consensus Definitions for Sepsis and Septic Shock (Sepsis-3). Jama 2016, 315(8):775–787.

20. Thompson BT, Chambers RC, Liu KD: Acute Respiratory Distress Syndrome. N Engl J Med 2017, 377(6):562–572.

21. Tamakuma S, Ogawa M, Aikawa N, Kubota T, Hirasawa H, Ishizaka A, Taenaka N, Hamada C, Matsuoka S, Abiru T: Relationship between neutrophil elastase and acute lung injury in humans. Pulm Pharmacol Ther 2004, 17(5):271–279.

22. Zeiher BG, Artigas A, Vincent JL, Dmitrienko A, Jackson K, Thompson BT, Bernard G: Neutrophil elastase inhibition in acute lung injury: results of the STRIVE study. Crit Care Med 2004, 32(8):1695–1702.

23. Aikawa N, Kawasaki Y: Clinical utility of the neutrophil elastase inhibitor sivelestat for the treatment of acute respiratory distress syndrome. Ther Clin Risk Manag 2014, 10:621–629.

24. Mohamed MMA, El-Shimy IA, Hadi MA: Neutrophil Elastase Inhibitors: A potential prophylactic treatment option for SARS-CoV-2-induced respiratory complications? Crit Care 2020, 24(1):311.

25. Denning NL, Aziz M, Gurien SD, Wang P: DAMPs and NETs in Sepsis. Front Immunol 2019, 10:2536.

26. Fei Y, Huang X, Ning F, Qian T, Cui J, Wang X, Huang X: NETs induce ferroptosis of endothelial cells in LPS-ALI through SDC-1/HS and downstream pathways. Biomed Pharmacother 2024, 175:116621.

27. Matsuzaki K, Hiramatsu Y, Homma S, Sato S, Shigeta O, Sakakibara Y: Sivelestat reduces inflammatory mediators and preserves neutrophil deformability during simulated extracorporeal circulation. Ann Thorac Surg 2005, 80(2):611–617.

28. Pan T, Tuoerxun T, Chen X, Yang CJ, Jiang CY, Zhu YF, Li ZS, Jiang XY, Zhang HT, Zhang H et al: The neutrophil elastase inhibitor, sivelestat, attenuates acute lung injury in patients with cardiopulmonary bypass. Front Immunol 2023, 14:1082830.

29. Li J, Qi Z, Li D, Huang X, Qi B, Feng J, Qu J, Wang X: Alveolar epithelial glycocalyx shedding aggravates the epithelial barrier and disrupts epithelial tight junctions in acute respiratory distress syndrome. Biomed Pharmacother 2021, 133:111026.

30. Liu XY, Xu HX, Li JK, Zhang D, Ma XH, Huang LN, Lü JH, Wang XZ: Neferine Protects Endothelial Glycocalyx via Mitochondrial ROS in Lipopolysaccharide-Induced Acute Respiratory Distress Syndrome. Front Physiol 2018, 9:102.

